# Oral and Maxillofacial Surgeon Accuracy in Anticipating Supplemental Opioid Use Following Third Molar Extraction

**DOI:** 10.64898/2026.07.02.26357136

**Authors:** Sierra R. van den Dries, Neeraj Panchal, Steven Wang, Rania A. Habib, Brian P. Ford, Stacey A. Secreto, Elliot V. Hersh, Katherine N. Theken

**Author notes:** **Corresponding Author**, Katherine N. Theken, PharmD, PhD, University of Pennsylvania School of Dental Medicine, Department of Oral and Maxillofacial Surgery and Pharmacology, 240 S 40th St, Room 531 Levy, Philadelphia, PA 19104.

## Abstract

**Background:** Accurately identifying patients who will require opioids after third molar extraction could improve pain management while supporting opioid stewardship. This study evaluated surgeon accuracy in predicting supplemental opioid use following treatment with ibuprofen and acetaminophen.

**Methods:** Patients (N=85) undergoing third molar extraction were treated with a standardized analgesic regimen of ibuprofen+acetaminophen, with supplemental opioid if needed. Four surgeons independently reviewed preoperative radiographs, assessed surgical difficulty using the Pederson scale, and rated the likelihood of supplemental opioid use on a 5-point Likert scale. Inter-rater reliability was assessed using intraclass correlation coefficients (ICC). The relationship between surgeon ratings and postoperative opioid use was evaluated using logistic regression and receiver operating characteristic (ROC) analysis.

**Results:** Seventeen patients used supplemental opioid analgesics. Inter-rater reliability among surgeons was moderate (ICC3=0.606, 95%CI: 0.505-0.700), while reliability of the average rating across surgeons was good (ICC3k = 0.860, 95% CI: 0.804-0.903). Median surgeon rating was not associated with postoperative opioid use (OR: 0.800, 95% CI: 0.414-1.51, p=0.496) and demonstrated poor discrimination (AUC: 0.551, 95% CI: 0.392-0.710). Surgeon ratings were positively associated with Pederson score (β=0.073, 95%CI: 0.050-0.096; p<0.001).

**Conclusions:** Surgeons demonstrated moderate agreement, but these assessments did not accurately identify patients who ultimately required supplemental opioids. Surgeon judgments appeared to be influenced by anticipated surgical difficulty.

**Practical Implications:** Clinicians should follow current recommendations against routine “just-in-case” opioid prescribing after third molar extraction. Future studies should focus on identifying clinical and biological predictors of inadequate analgesic response to NSAIDs to support individualized pain management strategies.

## Introduction

Non-steroidal anti-inflammatory drugs (NSAIDs) are recommended as first-line analgesics for most patients following third molar extraction.^1, 2^ These recommendations are based upon numerous clinical studies demonstrating that NSAIDs are as effective, and in some cases more effective, than opioid analgesics in managing acute dental pain.^3, 4^ However, there is variability in the analgesic response to NSAIDs, with several studies reporting that 20-30% patients required opioid rescue medication in addition to NSAID for adequate pain management.^5-9^

Post-operative pain management is a significant contributor to patient satisfaction, so dentists strive to anticipate patients’ needs prior to the procedure.10 However, providing adequate pain control must be balanced with the need to limit unnecessary opioid prescriptions. Often, a person is first exposed to opioids with extraction of third molars in adolescence or early adulthood. Even short term exposure to these drugs has been demonstrated to significantly increase the risk of subsequent opioid misuse and abuse.^11^ Concerningly, in Maughan et al, it was found that 54% of opioids prescribed by dentists for surgical procedures went unused.^12^ Eighty percent of high school students reporting medical use of opioids prior to misuse indicated sourcing the substance from their own previous prescription.^13^ In a study of almost 50,000 12^th^ graders from 2001-2005, 10-11% of these students indicated that they had experimented with hydrocodone/acetaminophen (Vicodin) in a recreational manner.^14^ These statistics emphasize the onus on providers to reduce exposure and circulation of opioids, specifically in the susceptible populations that present for third molar extractions.

Surgical difficulty and procedure length are often considered when deciding whether to prescribe opioids. Prior studies have reported that greater surgical difficulty, as assessed by tooth position and morphology, a 4-class rating scale (I, extraction requiring forceps only; II, extraction requiring osteotomy; III, extraction requiring osteotomy and coronal section; IV, complex extraction (root section)), and/or length of surgery, was associated with greater pain intensity following third molar extraction.^15-19^ However, no studies investigated whether surgical difficulty impacted the degree of pain relief subjects experienced or need for supplemental opioids when treated with evidence-based analgesia.

The goal of this study was to determine how well oral and maxillofacial surgeons could predict which patients would require supplemental opioids, in addition to ibuprofen and acetaminophen, for adequate pain management after third molar extraction based on pre-operative panoramic radiographs.

## Methods

This was a secondary analysis of data collected in a randomized, double-blind, placebo-controlled clinical study of subjects undergoing impacted third molar extraction. The study consisted of two phases: a double-blind, placebo-controlled inpatient phase of the analgesic response to ibuprofen in the acute post-surgical period and an open-label outpatient phase in which all subjects received the combination of ibuprofen and acetaminophen as their primary analgesic regimen. The primary endpoint was use of opioid rescue medication, in addition to ibuprofen and acetaminophen, in the first seven days following extraction. The study protocol was approved by the University of Pennsylvania Institutional Review Board (IRB#832417; ClinicalTrials.gov: NCT03893175), and all participants provided informed consent. The study was conducted in accordance with the Declaration of Helsinki.

Details of the study design and enrollment criteria have been reported previously.^9^ Briefly, healthy volunteers (≥18 years of age) were recruited from patients referred to the Oral and Maxillofacial Surgery Service at the School of Dental Medicine and the Hospital of the University of Pennsylvania for extraction of one or more partially or fully bony impacted mandibular third molar between May 2019 and March 2022. Maxillary third molars were included in the surgical plan, if appropriate.

Third molar extraction was performed by one of four attending oral surgeons (N.P.; S.W.; R.A.H; and B.P.F.) using a standardized regimen of 2% lidocaine plus 1:100,000 epinephrine for local anesthesia and nitrous oxide/oxygen or midazolam + fentanyl (maximum 50 mcg) titrated to effect for sedation to ensure adequate pain management during the procedure and relatively rapid dissipation of the effect once surgery was complete. After surgery participants reported pain intensity every 15 minutes using the 0-10 Numeric Rating Scale (NRS-PI), where 0=no pain and 10=worst pain imaginable. When study participants reported a pain score ≥4/10 or indicated that their pain was no longer tolerable, they received a blinded dose of rapid-acting ibuprofen liquigel 400 mg or matching placebo by mouth, randomized in a ratio of 3:1. Oxycodone 5mg was available as a rescue analgesic if pain relief was inadequate with the blinded study medication.

Four hours after blinded study medication treatment, all participants received open-label ibuprofen 400 mg and acetaminophen 500 mg to be taken every 4 h around the clock for the first 2 days and then as needed for pain.^1^ Additionally, participants were given 8 oxycodone 5 mg tablets to be taken up to every 6 h as needed in case they experienced insufficient pain relief on ibuprofen and acetaminophen. Outpatient analgesic use was recorded on a pain diary, which was returned on Day 7 after extraction with any unused study medication. Study data were collected and managed using REDCap electronic data capture tools hosted at the University of Pennsylvania Perelman School of Medicine.^20, 21^

### Evaluation of Surgical Difficulty and Likelihood of Supplemental Opioid Use

Panoramic radiographs were obtained in anticipation of subjects undergoing third molar surgery in a clinical setting. Four attending oral and maxillofacial surgeons (N.P.; S.W.; R.A.H; and B.P.F.) reviewed each patient’s panoramic radiograph to determine surgical difficulty for each molar based on the Pederson scale.^22^ Mandibular third molar difficulty was scored based on depth, angulation of impaction, and relationship to the ramus. Maxillary third molar difficulty was scored based on depth and angulation of impaction. The median of the oral surgeons’ scores for each molar was used for subsequent analysis. The resultant Pederson scores for each extracted tooth were summed to obtain an overall surgical difficulty for each patient. Based on the radiograph, surgeons also rated whether the patient would require supplemental opioids in addition to the scheduled ibuprofen and acetaminophen after extraction using a 5-point Likert scale (strongly disagree, disagree, neutral, agree, strongly agree).

### Statistical analysis

Data are reported as mean±standard deviation, median (interquartile range (IQR)), or percentages, as applicable. The surgeons’ rating of likelihood of supplemental opioid use was converted to an ordinal scale (1=strongly disagree, 2=disagree, 3=neutral, 4=agree, 5=strongly agree). Inter-rater reliability among the four surgeons was assessed using the intraclass correlation coefficient (ICC) derived from a two-way mixed-effects model for absolute agreement. The relationship between median surgeon rating and supplemental opioid use was evaluated by logistic regression, and discrimination was assessed using area under the receiver operating characteristic (ROC) curve. The relationship between summed Pederson score and surgeon rating was evaluated by Spearman correlation and linear mixed-effects modeling with summed Pederson score as a fixed effect and surgeon as a random effect. P<0.05 was considered statistically significant. Statistical analyses were performed in R (version 4.4.2), and figures were created in Graphpad Prism.

## Results

Eighty-seven healthy adults were enrolled. Two patients did not experience moderate pain in the first 4 hours after third molar extraction, and a panoramic radiograph was not available for one patient who did not use supplemental opioid, so these patients were excluded from further analysis. Thus, 84 patients were included in this study cohort (Table 1). Of these, 17 patients used supplemental oxycodone, while 67 used only ibuprofen+ acetaminophen for pain management.

**Table 1:**
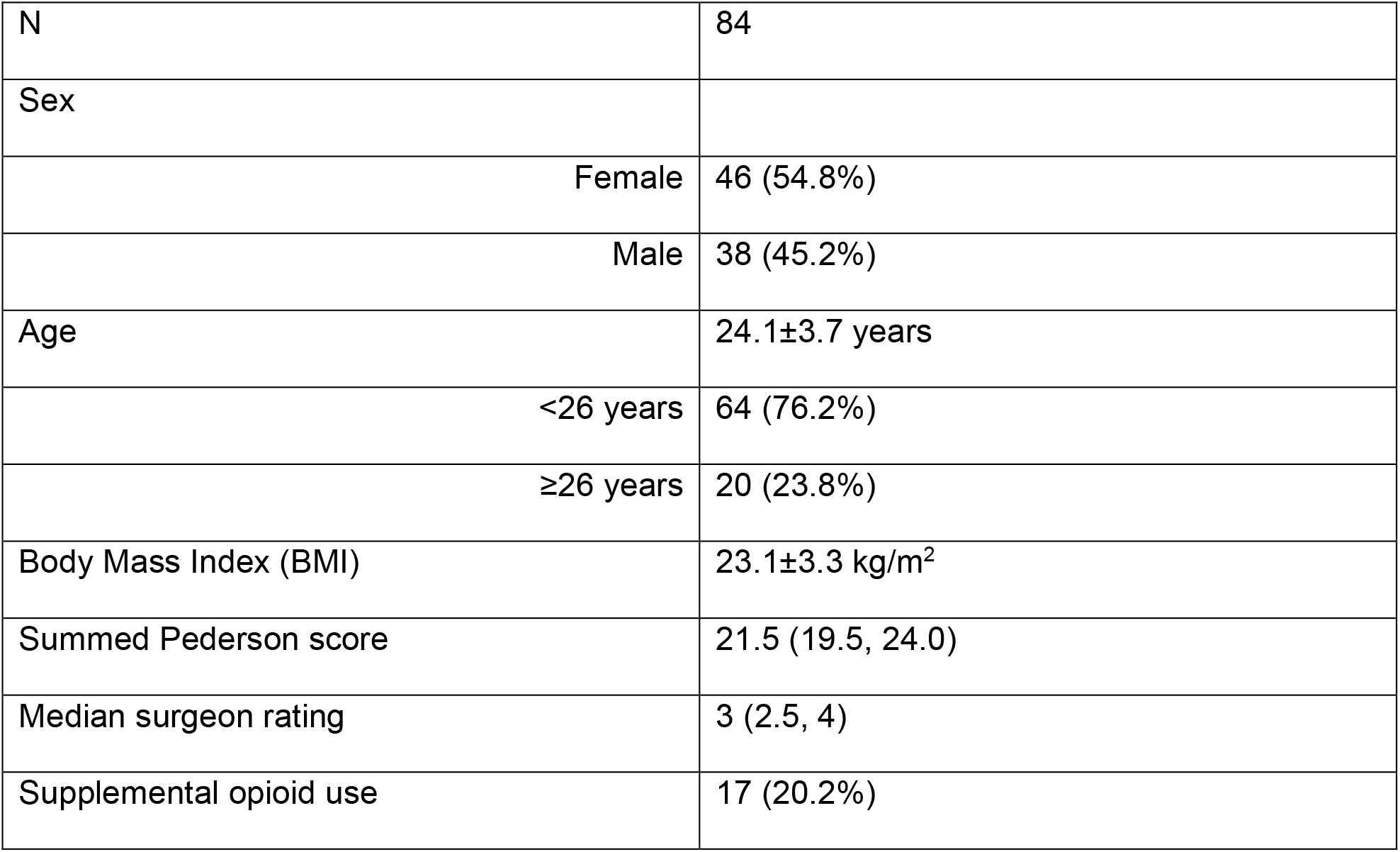
Cohort Characteristics.

Each surgeon’s prediction of whether a subject would require supplemental opioid use for pain control is shown in Figure 1. Inter-rater agreement among surgeons was moderate (ICC3=0.606, 95%CI: 0.505-0.700), while the agreement for the average rating across all four surgeons was good (ICC3k = 0.860, 95% CI: 0.804-0.903).

**Figure 1:**
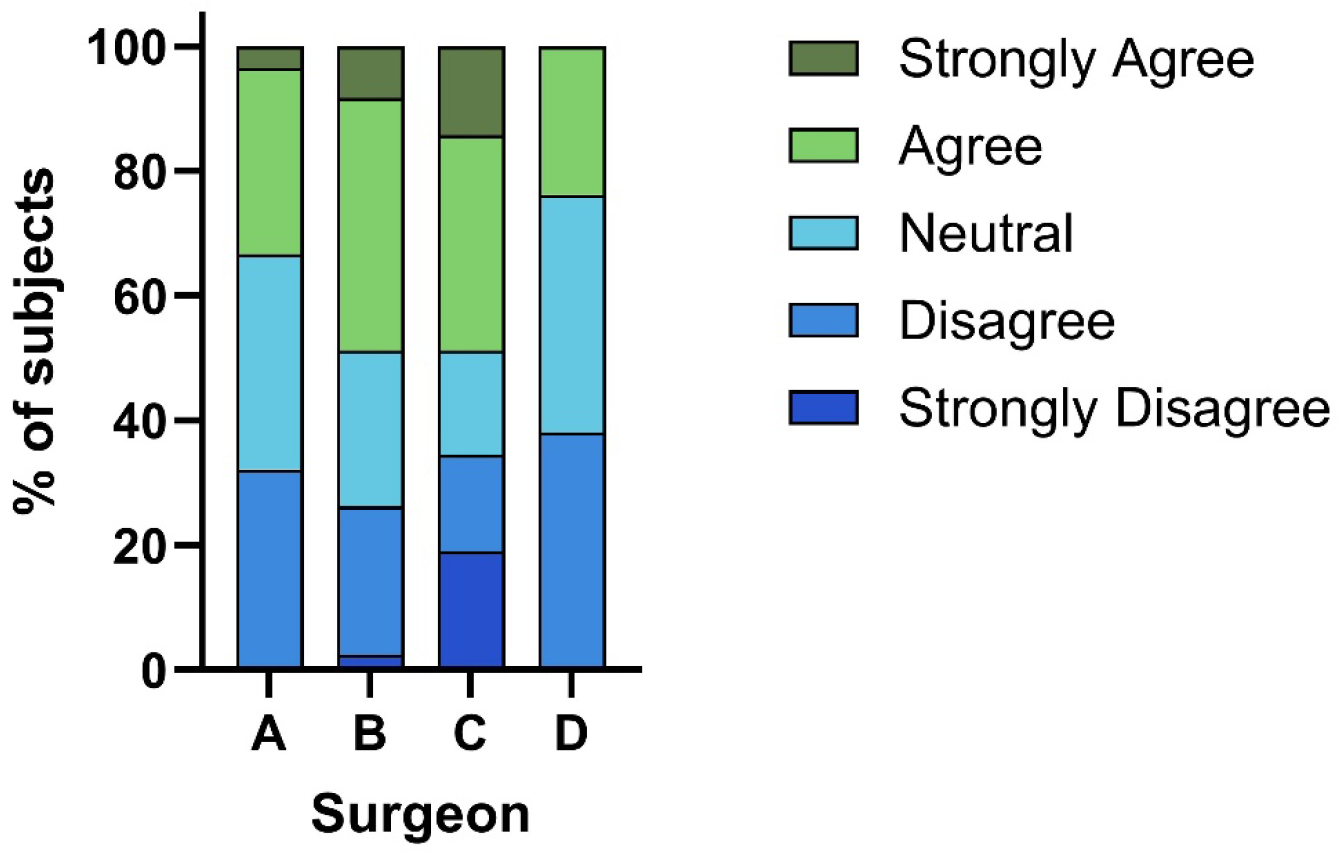
Individual surgeon ratings of likelihood of supplemental opioid use following third molar extraction.

Median surgeon rating was not associated with supplemental opioid use (OR: 0.800, 95% CI 0.414-1.51, p=0.496), and the surgeon ratings were similar between patients who used supplemental opioids (3 (2, 3.75) and those who did not (3 (2.5, 4), p=0.512; Figure 2A). The logistic regression model evaluating the association between surgeon rating and supplemental opioid use exhibited poor discrimination (AUC: 0.551, 95% CI: 0.392-0.710; Figure 2B).

**Figure 2:**
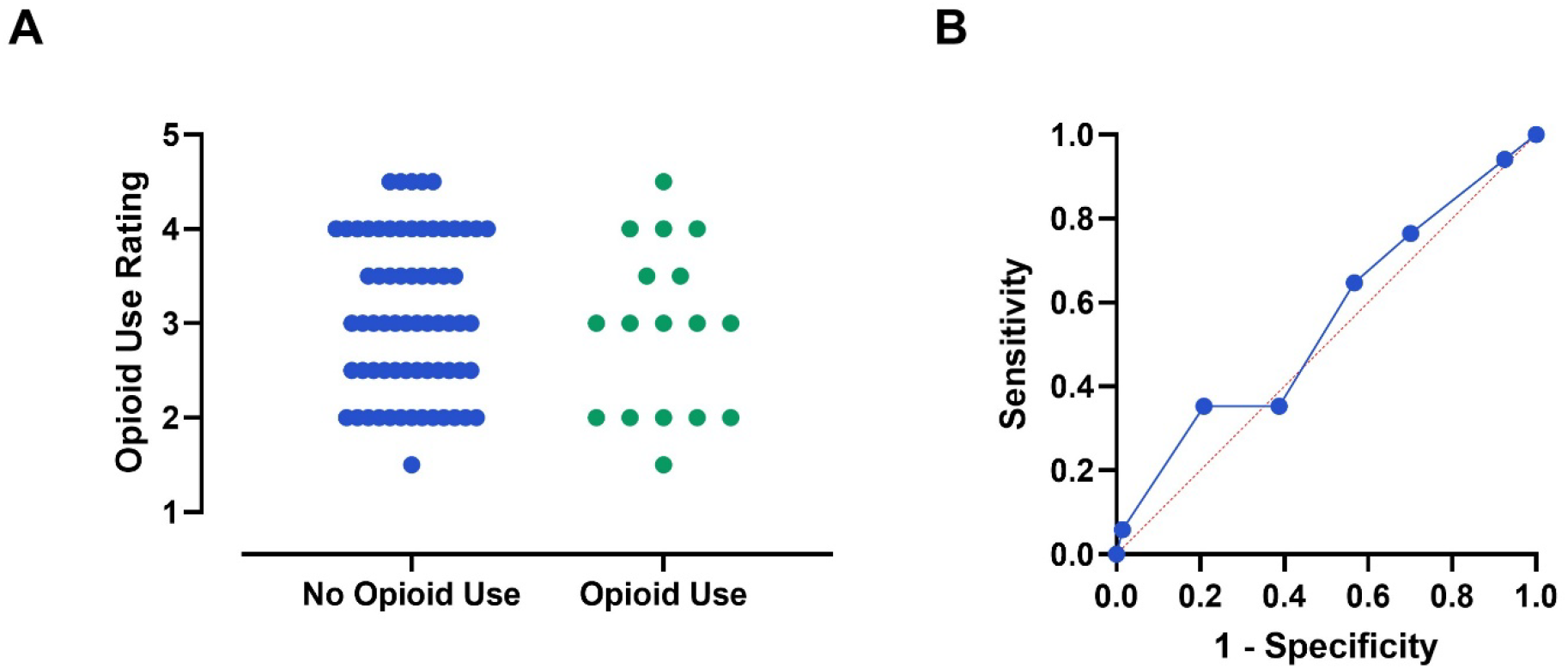
A) Distribution of median surgeon ratings by supplemental opioid use. B) Receiver operating characteristic (ROC) curve evaluating surgeon rating as a predictor of supplemental opioid use.

There was a moderate positive correlation between median surgeon rating of likelihood of supplemental opioid use and summed Pederson scores (Figure 3; ρ=0.344; p=0.001). Consistent with this finding, a linear mixed-effects model accounting for surgeon-level clustering indicated that the summed Pederson score was positively associated with surgeon rating (β=0.073, 95%CI: 0.050-0.096; p<0.001).

**Figure 3:**
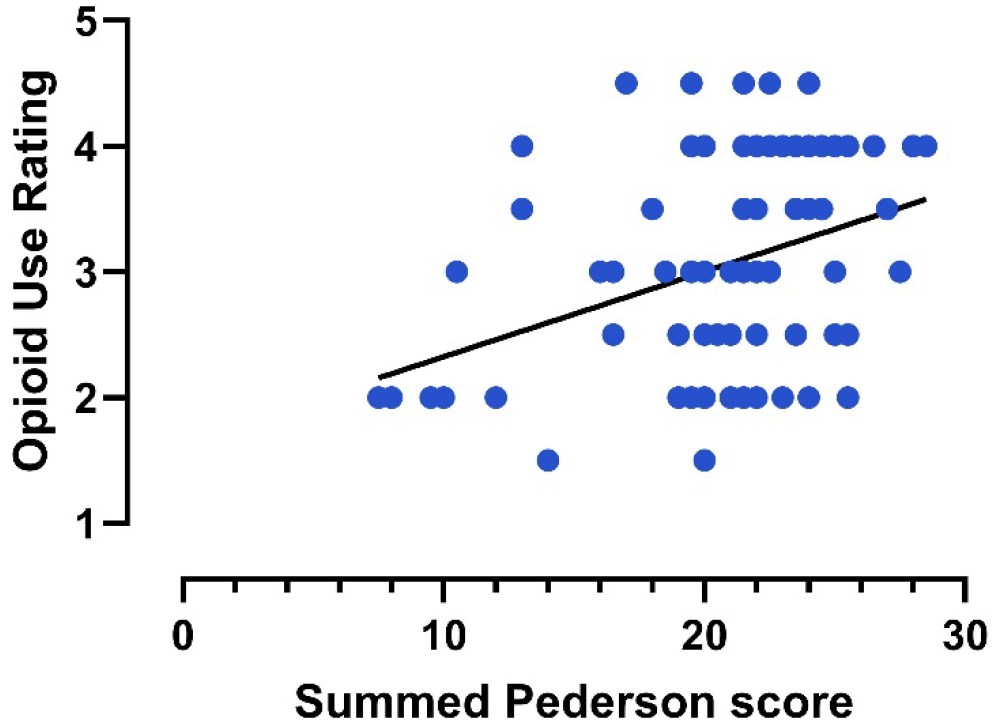
Relationship between summed Pederson score and median surgeon rating.

## Discussion

Our results indicate that oral and maxillofacial surgeons cannot accurately predict which patients will require supplemental opioids when treated with an evidence-based analgesic regimen following third molar extraction. Notably, we observed that surgeons tend to expect more patients to require opioids, in addition to ibuprofen + acetaminophen, in the outpatient period than they truly do. Surgeon ratings of likelihood of supplemental opioid use were associated with Pederson score, suggesting that clinical judgments regarding pain management may be influenced by anticipated surgical difficulty.

We observed a moderate degree of agreement among the surgeons, as evidenced by ICC=0.601. However, their assessments lacked predictive validity, with similar surgeon ratings between patients who used supplemental opioids and those who did not and poor discrimination in the logistic regression model. Importantly, the moderate inter-rater reliability suggests that the inability to predict opioid use was not primarily due to disagreement among surgeons.

Rather, surgeons appeared to rely on similar clinical cues that were not informative for identifying patients who ultimately required supplemental opioids. The surgeons also tended to overestimate the number of patients who would require supplemental opioids, with the proportion of Agree/Strongly Agree ratings ranging from 23.8% to 48.8%. This is consistent with the results of Maughan, et al. who showed that oral surgeons prescribed nearly twice as many opioid pills as patients actually used.^12^

The association between Pederson score and surgeon rating suggests that surgeons are considering surgical difficulty when evaluating whether or not a patient will require supplemental opioids. Several studies indicate that greater surgical difficulty, defined either based on radiographic assessment or by length of surgery, is associated with greater pain intensity following third molar extraction. ^15-19, 23, 24^ Thus, it is reasonable for clinicians to expect that patients undergoing more difficult surgical procedures will experience greater postoperative pain. However, the need for supplemental opioid analgesia depends not only on pain intensity but also on the effectiveness of first-line NSAID-based analgesia. Our prior work demonstrated that analgesic response to ibuprofen was associated with the degree of activation of the cyclooxygenase pathway, the molecular target of NSAIDs.^8^ Consequently, NSAIDs may provide excellent analgesia even in patients experiencing substantial postoperative pain when prostaglandin-mediated inflammation is a key driver. Conversely, patients with relatively modest pain intensity may experience inadequate analgesia if prostaglandin signaling contributes minimally to their pain. As a result, factors associated with anticipated pain intensity may not necessarily predict postoperative opioid use.

Our results underscore the importance of prescribing opioids in accordance with the clinical practice guidelines for acute dental pain.^1^ Numerous clinical studies support the safety and efficacy of NSAIDs in managing acute dental pain, ^3, 4^ and most patients can be effectively managed with NSAID-based regimens alone.^8, 9^ Surgeons cannot reliably identify which patients may require supplemental opioid preoperatively. Thus, routine “just-in-case” opioid prescriptions are difficult to justify, and supplemental opioid prescriptions should be reserved for patients reporting inadequate analgesia with NSAIDs.

This study has several strengths. Patients were managed within a prospective clinical study with standardized postoperative analgesic regimens, which minimized provider bias in providing an opioid prescription. Further, by promoting high adherence to first-line NSAID therapy, the study design reduced variability attributable to medication-taking behavior and enabled a more direct assessment of factors associated with inadequate analgesic response. In addition, we had detailed assessment of medication use, allowing for robust ascertainment of supplemental opioid use. All patients were independently evaluated by four oral surgeons, enabling assessment of inter-rater reliability.

There are also several limitations to our study. Because participants were enrolled in a clinical research study, patient behavior and medication use may not fully reflect routine clinical practice. In addition, the relatively small number of patients who used supplemental opioids (N=17) limited our statistical power to detect modest predictive effects and precluded subgroup analyses. Thus, we were unable to determine whether surgeons were better able to predict need for supplemental opioid in different age or sex groups. In particular, patients ≥26 years of age are at higher risk of complications following third molar extraction.^2^ All surgeons involved in the study operate in an academic center. Because these surgeons are often treating patients with complex oral and maxillofacial conditions, perhaps they are primed to anticipate a need for opioids more readily than a private practice surgeon might. Additionally, surgeons based their assessments on preoperative radiographs and limited clinical information, which may not capture all factors that are considered when prescribing opioids in a clinical setting.

## Conclusions

In summary, surgeons demonstrated moderate agreement in predicting post-surgical opioid use in patients treating with an evidence-based analgesic regimen following third molar extraction, but these assessments did not accurately identify patients who ultimately used supplemental opioids. These findings suggest that clinical factors commonly used to anticipate postoperative pain and opioid requirements, such as surgical difficulty, may not adequately capture the determinants of analgesic response to NSAIDs. Thus, clinicians should follow current recommendations against routine “just-in-case” opioid prescribing following third molar extraction. Future research elucidating the clinical and biological predictors of inadequate analgesic response to NSAIDs may enable more individualized and evidence-based pain management strategies.

## Data Availability

All data produced in the present study are available upon reasonable request to the authors

## Acknowledgements

This work was supported by an Oral and Maxillofacial Surgery Foundation Clinical Research Grant and funding from the National Institute of Dental and Craniofacial Research (R01DE033405). The content is solely the responsibility of the authors and does not necessarily represent the official views of the National Institutes of Health. The funders had no role in the study design, in the collection, analysis and interpretation of data; in the writing of the manuscript; or in the decision to submit the manuscript for publication. The authors have no conflicts of interest to disclose.

## Notes

### Competing Interest Statement

The authors have declared no competing interest.

### Author Declarations

The Institutional Review Board of the University of Pennsylvania gave ethical approval for this work.

## References

1. Carrasco-Labra A, Polk DE, Urquhart O, et al. Evidence-based clinical practice guideline for the pharmacologic management of acute dental pain in adolescents, adults, and older adults: A report from the American Dental Association Science and Research Institute, the University of Pittsburgh, and the University of Pennsylvania. J Am Dent Assoc 2024;155(2):102–17 e9.

2. American Association of Oral and Maxillofacial Surgeons Opioid Prescribing: Acute and Postoperative Pain Management. 2017. “https://www.aaoms.org/docs/govt_affairs/advocacy_white_papers/opioid_prescribing.pdf“. Accessed April 30 2020.

3. Miroshnychenko A, Ibrahim S, Azab M, et al. Acute Postoperative Pain Due to Dental Extraction in the Adult Population: A Systematic Review and Network Meta-analysis. J Dent Res 2023;102(4):391–401.

4. Hersh EV, Moore PA, Grosser T, et al. Nonsteroidal Anti-Inflammatory Drugs and Opioids in Postsurgical Dental Pain. J Dent Res 2020;99(7):777–86.

5. Dionne RA, Campbell RA, Cooper SA, Hall DL, Buckingham B. Suppression of postoperative pain by preoperative administration of ibuprofen in comparison to placebo, acetaminophen, and acetaminophen plus codeine. J Clin Pharmacol 1983;23(1):37–43.

6. Hersh EV, Levin LM, Cooper SA, et al. Ibuprofen liquigel for oral surgery pain. Clin Ther 2000;22(11):1306–18.

7. Hersh EV, Levin LM, Adamson D, et al. Dose-ranging analgesic study of Prosorb diclofenac potassium in postsurgical dental pain. Clin Ther 2004;26(8):1215–27.

8. Theken KN, Hersh EV, Lahens NF, et al. Variability in the Analgesic Response to Ibuprofen Is Associated With Cyclooxygenase Activation in Inflammatory Pain. Clin Pharmacol Ther 2019;106(3):632–41.

9. Panchal N, Wang S, Habib RA, et al. Predictors of Supplemental Opioid Use After Third Molar Extraction. medRxiv 2025.

10. Berkowitz R, Vu J, Brummett C, et al. The Impact of Complications and Pain on Patient Satisfaction. Ann Surg 2021;273(6):1127–34.

11. Schroeder AR, Dehghan M, Newman TB, Bentley JP, Park KT. Association of Opioid Prescriptions From Dental Clinicians for US Adolescents and Young Adults With Subsequent Opioid Use and Abuse. JAMA Intern Med 2019;179(2):145–52.

12. Maughan BC, Hersh EV, Shofer FS, et al. Unused opioid analgesics and drug disposal following outpatient dental surgery: A randomized controlled trial. Drug Alcohol Depend 2016;168:328–34.

13. McCabe SE, West BT, Teter CJ, Boyd CJ. Medical and nonmedical use of prescription opioids among high school seniors in the United States. Arch Pediatr Adolesc Med 2012;166(9):797–802.

14. Friedman RA. The changing face of teenage drug abuse--the trend toward prescription drugs. N Engl J Med 2006;354(14):1448–50.

15. Bortoluzzi MC, Guollo A, Capella DL. Pain levels after third molar surgical removal: an evaluation of predictive variables. J Contemp Dent Pract 2011;12(4):239–44.

16. de Santana-Santos T, de Souza-Santos a A, Martins-Filho PR, et al. Prediction of postoperative facial swelling, pain and trismus following third molar surgery based on preoperative variables. Med Oral Patol Oral Cir Bucal 2013;18(1):e65–70.

17. Grossi GB, Maiorana C, Garramone RA, et al. Assessing postoperative discomfort after third molar surgery: a prospective study. J Oral Maxillofac Surg 2007;65(5):901–17.

18. Lago-Mendez L, Diniz-Freitas M, Senra-Rivera C, et al. Relationships between surgical difficulty and postoperative pain in lower third molar extractions. J Oral Maxillofac Surg 2007;65(5):979–83.

19. Wang TF, Wu YT, Tseng CF, Chou C. Associations between dental anxiety and postoperative pain following extraction of horizontally impacted wisdom teeth: A prospective observational study. Medicine (Baltimore) 2017;96(47):e8665.

20. Harris PA, Taylor R, Minor BL, et al. The REDCap consortium: Building an international community of software platform partners. J Biomed Inform 2019;95:103208.

21. Harris PA, Taylor R, Thielke R, et al. Research electronic data capture (REDCap)--a metadata-driven methodology and workflow process for providing translational research informatics support. J Biomed Inform 2009;42(2):377–81.

22. Pedersen GW. Oral surgery. Philadelphia: Saunders; 1988.

23. Ali HT, Mosleh MI, Shawky M. Variables predictive of the intensity of postoperative pain following mandibular third molar surgery: a prospective study. Minerva Stomatol 2018;67(3):111–16.

24. Benediktsdottir IS, Wenzel A, Petersen JK, Hintze H. Mandibular third molar removal: risk indicators for extended operation time, postoperative pain, and complications. Oral Surg Oral Med Oral Pathol Oral Radiol Endod 2004;97(4):438–46.

